# Cluster analysis successfully identifies clinically meaningful knee valgus moment patterns: Frequency of early peaks reflects sex-specific ACL injury incidence

**DOI:** 10.1101/19001511

**Authors:** Haraldur B. Sigurðsson, Kristín Briem

## Abstract

**Background:** Biomechanical studies of ACL injury risk factors report heterogenous results, analyze only a fraction of the data, and do not analyze data in accordance with the injury mechanism. Extracting a peak value of relevance to ACL injuries is challenging due to differences in the relative timing of the peak value.

**Aims/Hypotheses:** The aim was to cluster analyze the knee valgus moment time series curve shape in the early stance phase. We hypothesized that 1a) There would be few discrete curve shapes, 1b) there would be an early peak pattern, 2a) youth athletes of both sexes would show similar frequencies of early peaks, 2b) adolescent girls would have higher early peak frequencies.

**Methods:** N=213 (39% boys) youth soccer and team handball athletes (phase 1) and N=35 (45% boys) with 5 year follow-up data (phase 2) were recorded performing a change of direction task with 3D motion analysis and a force plate. The time series of the first 30% of stance phase were cluster analyzed based on Euclidean distances in two steps; shape-based main clusters with a transformed time series, and magnitude based sub-clusters with body weight normalized time series. Group differences (sex, phase) in curve shape frequencies, and shape-magnitude frequencies were tested with chi-squared test.

**Results:** 6 discrete shape-clusters and 14 magnitude based sub-clusters were formed. Phase 1 boys had higher frequencies of early peaks than phase 1 girls (38% vs 25% respectively, P<0.001 for full test). Phase 2 girls had higher frequencies of early peaks than phase 2 boys (42% vs 21% respectively, P<0.001 for full test).

**Conclusions:** Cluster analysis can reveal different patterns of curve shapes in biomechanical data. The early peak shape is relatable to the ACL injury mechanism as the timing of its peak moment is consistent with the timing of injury. Higher frequency of early peaks demonstrated by Phase 2 girls is consistent with their higher incidence of ACL injury.

## Introduction

The biomechanical study of risk factors for anterior cruciate ligament (ACL) injury has revolved primarily around simplistic analyses of peak values extracted from a time series while ignoring much of the research on ACL injury mechanisms (Dai et al. 2014), including the likely timing of actual ACL rupture (Krosshaug et al. 2007). A landmark study by Hewett et al. (Hewett et al. 2005) revealed that the knee valgus moment was a risk factor for ACL injury. The study has been widely cited (more than 400 citations) despite some methodological limitations. The total number of injured players was low (N = 9), leading to a high chance of accidental results when dealing with highly variable data. Furthermore, the study used a bilateral drop-jump, a movement which typically does not result in athletic ACL injuries (Montgomery et al. 2018; Walden et al. 2015). Recent studies using similar methodology (Krosshaug et al. 2016; Leppanen et al. 2017) have not replicated the results of the Hewett study (Hewett et al. 2005) and the observation has been made that biomechanical risk factor studies do not account for the ACL loading mechanisms in their analyses (Dai et al. 2014) which may explain their inconsistent results.

The ACL injury mechanism is a complex multi-planar force event (Kiapour et al. 2014). While the peak knee valgus moment during a bilateral drop-jump is a poor predictor of ACL injuries, cadaveric studies have repeatedly demonstrated that the ACL can be loaded and potentially injured with a knee valgus moment in combination with other forces (Bates et al. 2018; Kiapour et al. 2016; Markolf et al. 1990). Considering that the timing of ACL rupture (Krosshaug et al. 2007) is inconsistent with the timing of the absolute *peak* knee valgus moment found during the weight acceptance phase of a dynamic task (Sigurðsson et al. 2018) it’s possible that the peak knee valgus moment is not a risk factor but rather the occurrence of an *early peak* knee valgus moment.

Since ACL injuries are rare (Montalvo et al. 2018) despite occurring during routine sports movements (Montgomery et al. 2018; Walden et al. 2015), the biomechanical risk factors for ACL injuries may also be observed rarely. Therefore, statistical tests of group differences that assess the early peak knee valgus moment magnitude alone are unlikely to identify true ‘at-risk’ individuals, because the majority of movement trials do not have a peak value with timing consistent with the injury (Krosshaug et al. 2007).

It’s been suggested that combining biomechanical research with machine learning tools, such as cluster analysis (Halilaj et al. 2018) may represent new avenues of research. Identifying a force-time curve shape consistent with the timing of ACL injury is a classification problem that may be solved with cluster analysis. To date, no method has been published that clusters force-time curves into different shapes.

The primary aim of this study was to use a cluster analysis technique to identify the different shapes of force-time curves of the knee valgus moment in the early weight acceptance phase of a change of direction task, a movement during which ACL injuries occur (Walden et al. 2015). Our hypotheses were; 1a) the force-time curves may be classified into a small number of different shapes using a cluster analysis algorithm, 1b) at least one of the resulting clusters will have a shape with the potential for a high knee valgus moment with timing consistent with ACL injury (Krosshaug et al. 2007).

A secondary aim was to compare the frequency of the early peak shape between the sexes before and after puberty. Our hypotheses were that; 2a) before adolescence, athletes will show an identical frequency of early peaks, 2b) after adolescence girls will have higher frequencies of early peaks, consistent with the 2-3x increased incidence reported in the literature (Montalvo et al. 2018; Nicholls et al. 2017).

## Methods

### Design and setting

Prospective cohort laboratory study.

### Subjects

Athletes (N=293) were 9-12 years old at study onset (phase 1) and recruited from local soccer and team handball clubs between 2012-2014. In order to have 120 participants at phase 2 despite the lower limit of estimated follow up of 40%, a sample size of 300 was required. This age range has been shown to have identical ACL injury rates (Nicholls et al. 2017) in the country where the study is performed. Participants signed an informed consent during first data collection that included permission to be contacted for the second phase. For the follow up, participants were contacted with the same contact information, and when required contact information from public records was sought. At the follow up data collection (phase 2), these same athletes (some of whom have changed, or departed from, sports) were aged 14-17 years old for a mean time from baseline to follow up of 5 years. In order to develop the methodology, a convenience sample of processed data from phase 1, and the currently available data from phase 2 were included. Both phase 1 athletes’ (N = 213, 39% boys) and phase 2 athletes (N = 35, 45% boys) characteristics are summarized in Table 1.

**Table 1:** Descriptive Statistics

|  |  | Boys |  | Girls |  | P |
| --- | --- | --- | --- | --- | --- | --- |
|  |  | Mean | SD | Mean | SD |  |
| Phase 1 | Age | 10.6 | 0.70 | 10.8 | 1.06 | 0.148 |
|  | Height | 149.0 | 7.87 | 149.9 | 8.20 | 0.721 |
|  | Weight | 40.2 | 8.10 | 41.8 | 9.38 | 0.402 |
|  | N | 1512 |  | 2502 |  |  |
| Phase 2 | Age | 15.8 | 0.81 | 16.0 | 0.77 | 0.500 |
|  | Height | 180.7 | 9.11 | 167.4 | 3.99 | < 0.001 |
|  | Weight | 74.9 | 16.54 | 64.0 | 10.27 | 0.054 |
|  | N | 364 |  | 419 |  |  |
| N is the total number of recorded movement trials per group. |  |  |  |  |  |  |

### Data collection

Data collection methods have been previously described by Briem et al (Briem et al. 2017). In short, height and weight were measured before a short warm-up on a stationary bike. Strength testing of hip muscles in abduction and external rotation was performed.

After strength testing, 46 reflective markers were placed on the subject, 4 on each foot, one per malleolus, a 4 marker cluster on each shank, one per femoral condyle, a 4 marker cluster on each thigh, a 3 marker cluster on the sacrum, one on each greater trochanter of the femur and on the highest point of each iliac crest, on bilateral anterior superior iliac spines, on the thorax (approximately t10-t12), on the c7, on the sternum, and on the lateral aspects of each scapular acromion.

A static trial was recorded, and anatomical markers were removed (trochanteric, malleolic, condylic, and iliac crests) before the dynamic movement trials. Subjects performed 5 repetitions of a change of direction task on each leg, and 5 repetitions of a bilateral drop-jump from a 23cm (youth) or 30cm (adolescents) box. Movement tasks were repeated after a 5 minute skateboard exercise protocol and all conditions were pooled for this analysis. The order of movement trials was randomized with an online randomizer in phase 2 (Haahr and Haahr), and a coinflip in phase 1.

### Data processing and statistical analysis

An 8 segment, 48 degree of freedom, musculoskeletal model was constructed in Visual3D (C-Motion) consisting of feet, shanks, thighs, a pelvis and a trunk. Ankle joint centers were defined as midway between malleolic markers, knee joint centers as midway between femoral condyle markers, hip joint centers as 25% of the distance between trochanteric markers, and the pelvis-trunk joint as midway between the iliac crest markers. Visual3D default settings were used for all segment inertial parameters.

Calculations of kinematics were performed using the 6 degree-of-freedom method and inverse kinetics were calculated for joint moments. Joint moments were normalized by subject body weight, since the tensile strength of the ACL ligament also scales with body weight (Chandrashekar et al. 2006). Time series data of the stance phase of a change of direction task was exported from Visual3D (C-Motion) and imported into R (Team 2018) for analysis. ACL injuries occur in the initial 50 ms after contact with the ground (Krosshaug et al. 2007) as revealed by video analysis of ACL injuries. Descriptions of ACL injuries most often involve high level athletes (Koga et al. 2018) due to the availability of match video recordings. We observed that the fastest athletes in our cohort who displayed a local peak knee valgus moment did so close to the 50 ms which generally was before 25% of the stance phase. To allow the curve to come down as well, a cut-off of 30% was selected.

### Cluster analysis

To measure the distance based on curve shapes, each time series was interpolated to lengths equal to the longest series + 2 and transformed by calculating the lagged differences of the series and taking its sign. A number that is higher than its previous number was equal to 1, and a lower number was equal to -1. Each time series was therefore reduced to its directional changes and therefore its scaled shape. The Euclidean distances between the transformed time series’ were calculated (Montero and Vilar 2014) and the distances were clustered using the Ward.D2 (Charrad et al. 2014; Murtagh and Legendre 2014) method which produces compact spherical clusters.

To decide on a number of clusters to produce, the C-Index (Hubert and Levin 1976) was calculated for clusters from 2:50. A number of clusters where the C-Index was below 0.05 was selected. The resulting clusters were examined and assigned a shape based on their appearance. Individual curves within the cluster were examined when the aggregated cluster appearance was unclear.

In order to differentiate different magnitudes of similar shapes, a second cluster analysis step was performed. All curves within each shape were interpolated and divided by bodyweight in kg. The Euclidean distances between them were calculated and using the Ward.D2 method (Murtagh and Legendre 2014), 2-4 sub-clusters based on force magnitude were formed. The lowest C-Index value out of the result was selected. Each of the resulting sub-clusters were then examined and classified as either a small, medium, or a large magnitude.

### Comparison between sexes

To determine differences in the frequency of each curve shape, a chi-square test was performed. The frequencies of each cluster shape for the categorical variables of phase and sex were calculated. The distribution of each cluster subgroup for maturity and sex were also calculated. Significance level was set at 0.05.

## Results

After screening for errors in performing the maneuver and large artifacts in the collected data, 4903 attempts out of the 5080 collected were available for analysis.

### Cluster analysis

After reducing each time series to the signs of a lagged difference, a total of 1025 unique shapes were present with a median of 1 trials per shape but with two large groups of identical shapes (Figure 1). A total of 39 clusters were formed in the initial cluster analysis step. No elbow was observed in the C-Index plot and the C-Index for 39 clusters was 0.049. From those 39 clusters, 6 distinct shapes were identified (Figure 2); early peaks, peaks, upslopes, downslopes, early troughs, and troughs. From the six basic shapes, a total of 14 magnitude based sub-clusters were formed (Figures 3 & 4).

**Figure 1:**
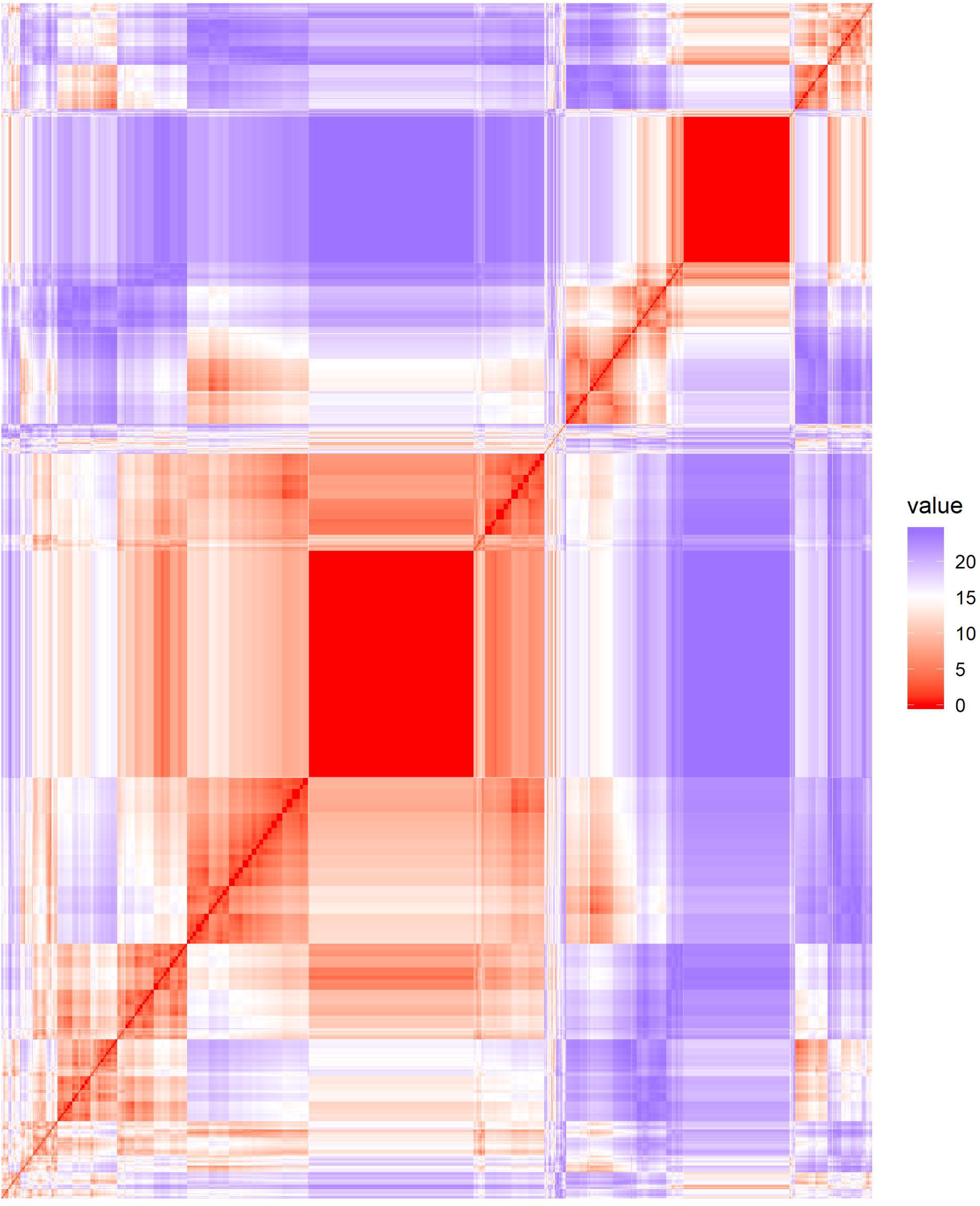
Heat map of the Euclidean distances of the time series after reduction to the signs of the differenced curve. Two large solid red boxes are present, indicating a number of identical time series (distance = 0).

**Figure 2:**
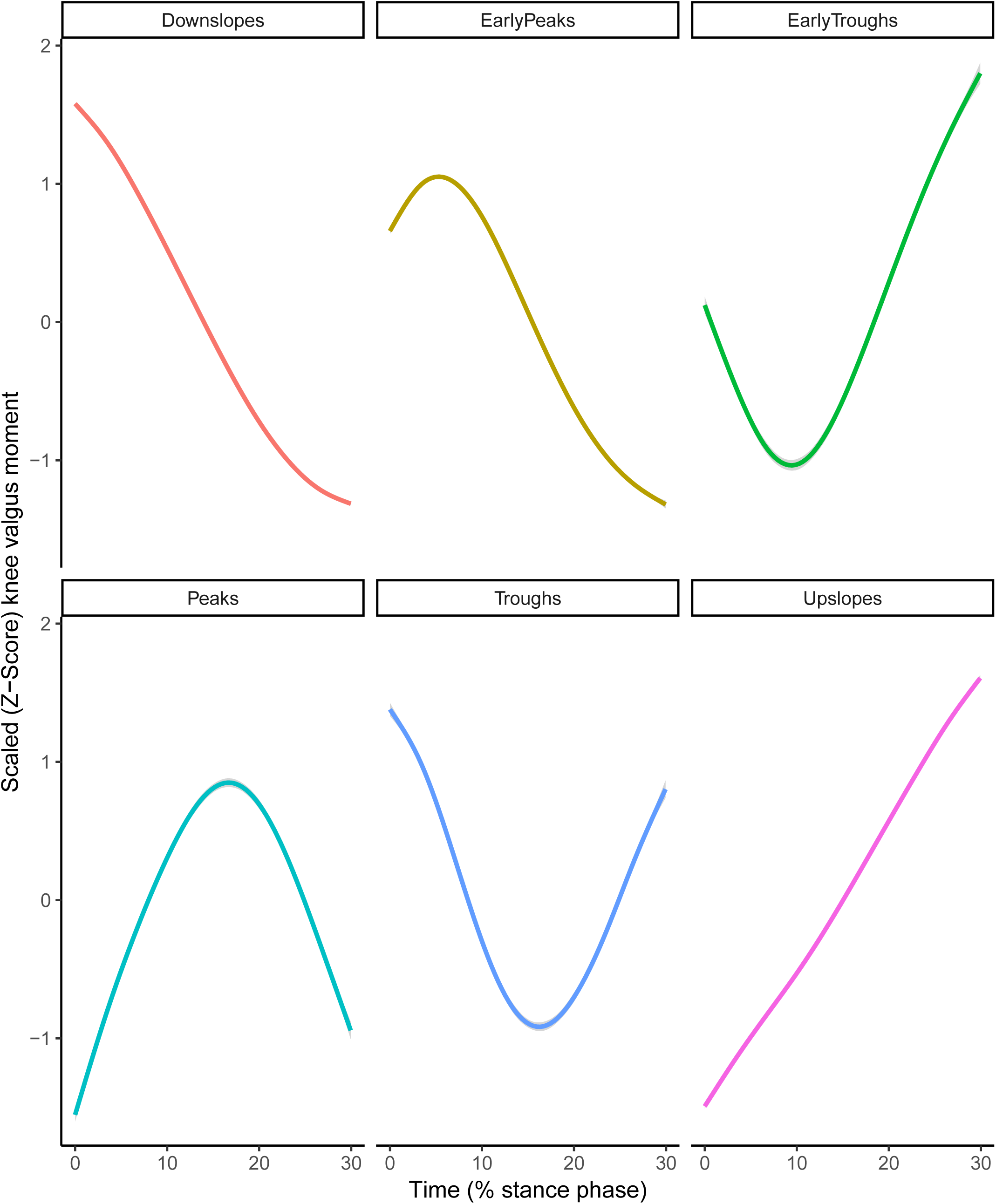
Smoothed aggregate time series’ of the six basic shapes of the scaled knee valgus moment curves generated in the initial shape-based cluster analysis. Each time series is individually scaled. The gray shaded area denotes the 95% confidence interval of the smoothing process.

**Figure 3:**
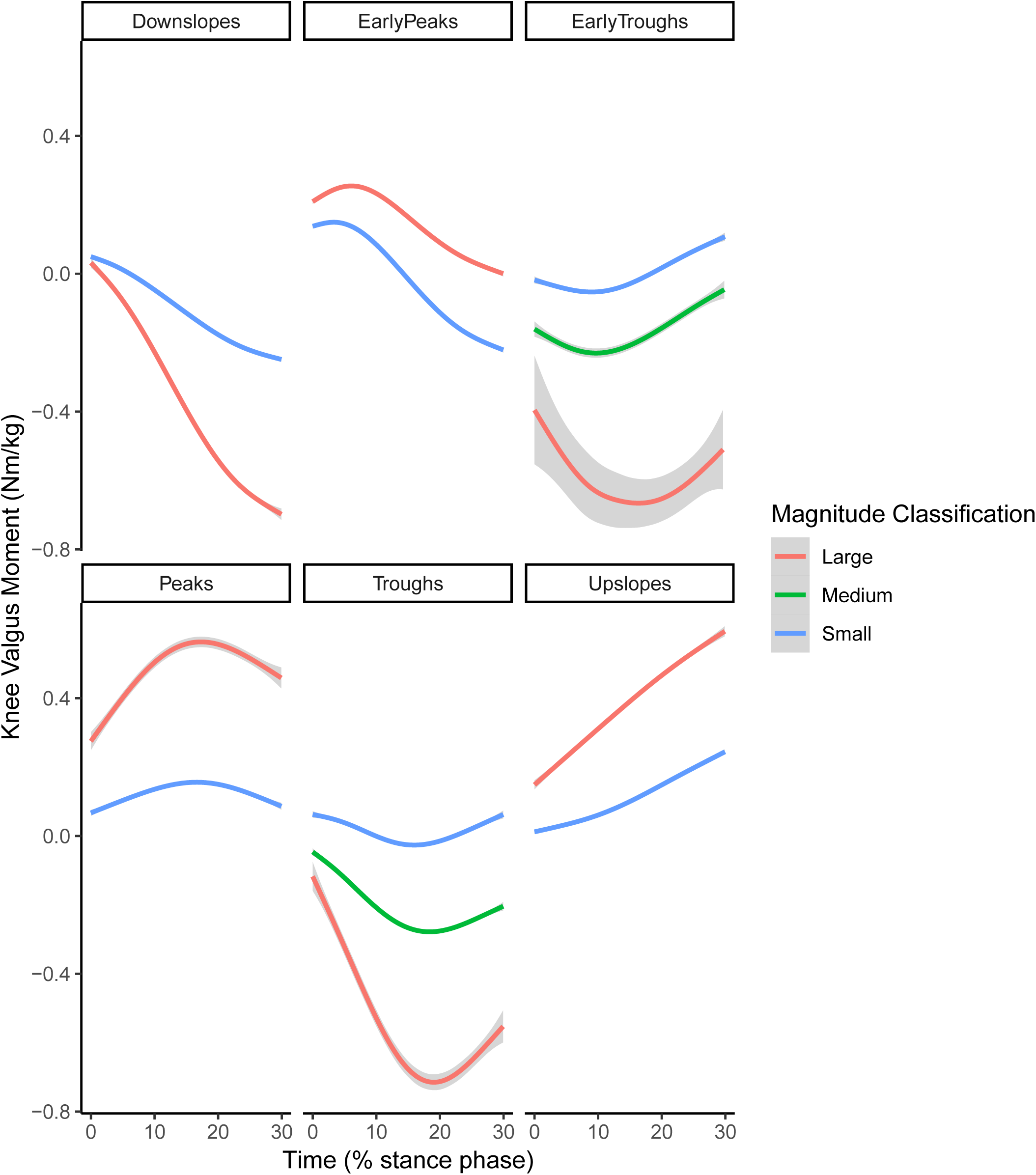
Smoothed aggregates of the time series of the first 30% of the stance phase of all clusters generated with the two step cluter analysis. The gray shaded area denotes the 95% confidence interval from the smoothing process.

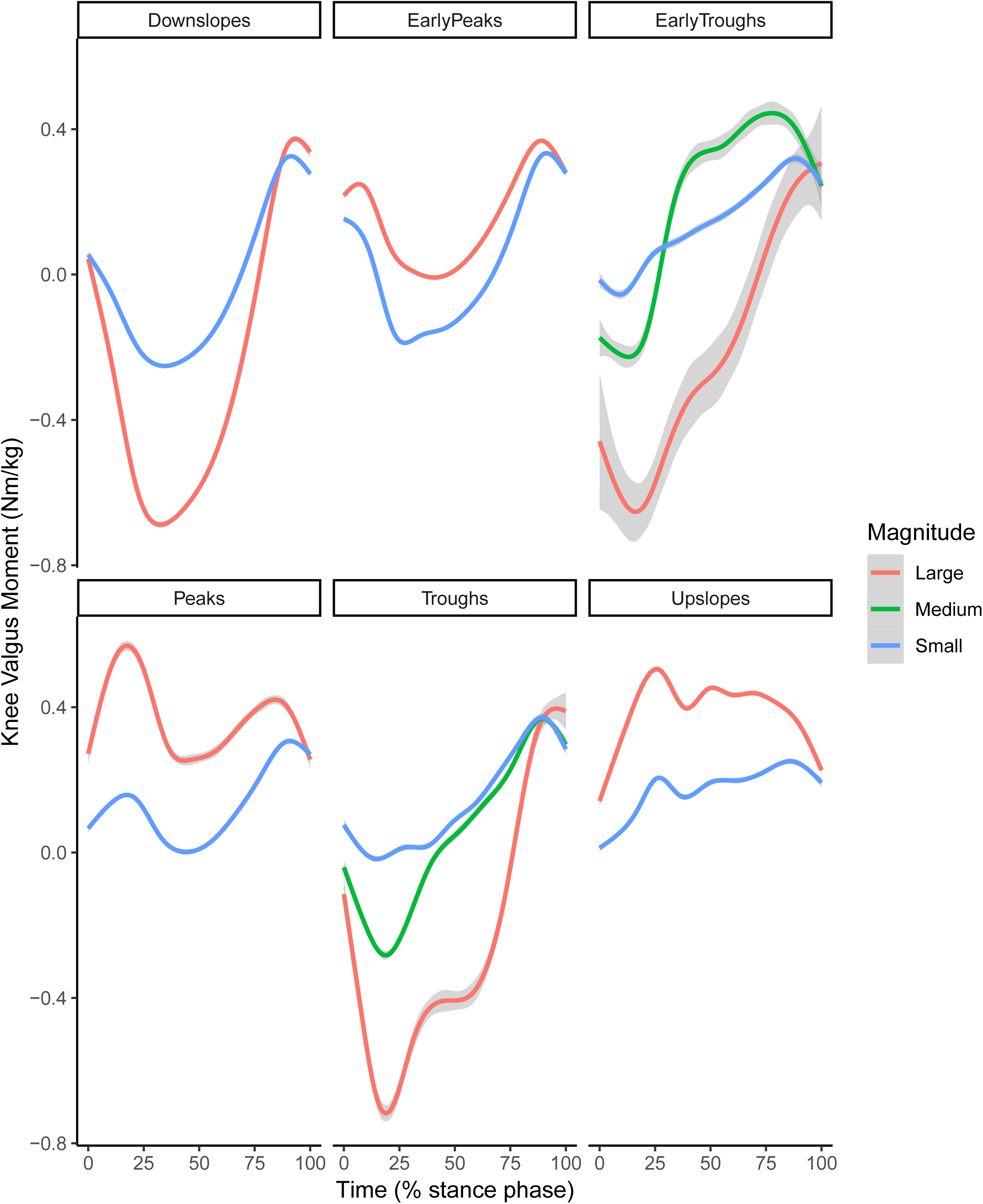

### Comparison between sexes

The chi-squared test for the six basic shapes revealed that in phase 1, boys had a higher than expected frequency of early peaks, while girls had a lower than expected frequency (chi-square contributions of 26.4 and 20.8, respectively). In phase 2, boys had a lower than expected frequency while girls had a higher than expected frequency of early peak shapes (chi-square contributions of 10.2 and 18.9, respectively). The total Chi-Square value of the test was 400.1 with P < 0.001. The frequencies, expected frequencies and chi-square contributions for shapes are reported in Table 2.

**Table 2:** The observed and expected frequencies of the six shape-based clusters

|  |  | Observed (Expected) |  | Chi-square contribution |  |
| --- | --- | --- | --- | --- | --- |
|  |  | Boys | Girls | Boys | Girls |
| Phase 1 | Downslopes | 276 (420) | 698 (695) | 49.46 | 0.01 |
|  | Early Peaks | 575 (464) | 642 (768) | 26.40 | 20.76 |
|  | Early Troughs | 42 (41) | 80 (67) | 0.04 | 2.40 |
|  | Peaks | 154 (149) | 272 (247) | 0.14 | 2.48 |
|  | Troughs | 160 (164) | 278 (272) | 0.11 | 0.14 |
|  | Upslopes | 305 (273) | 532 (452) | 3.68 | 14.08 |
| Phase 2 | Downslopes | 207 (101) | 152 (116) | 110.77 | 10.87 |
|  | Early Peaks | 78 (112) | 178 (129) | 10.20 | 18.92 |
|  | Early Troughs | 4 (10) | 3 (11) | 3.42 | 6.07 |
|  | Peaks | 10 (36) | 38 (41) | 18.75 | 0.28 |
|  | Troughs | 55 (40) | 28 (46) | 6.05 | 6.74 |
|  | Upslopes | 10 (66) | 20 (76) | 47.31 | 41.01 |
| Chi-square contribution is the individual cell contribution to Chi-square value from the chi square test. P-value for the Chi-Square test < 0.001 |  |  |  |  |  |

The relative frequency of the early peak shape overall was 32% in phase 1 and 32% in phase 2. The relative frequencies of the sexes were such that in phase 1 boys showed an early peak in 38% of trials while girls showed an early peak in 25% of trials. In phase 2 boys showed an early peak frequency of 21% (decrease from phase 1) while girls showed an early peak frequency of 42% (increase from phase 1). The relative frequency of each shape by sex and phase are shown in Figure 5.

**Figure 5:**
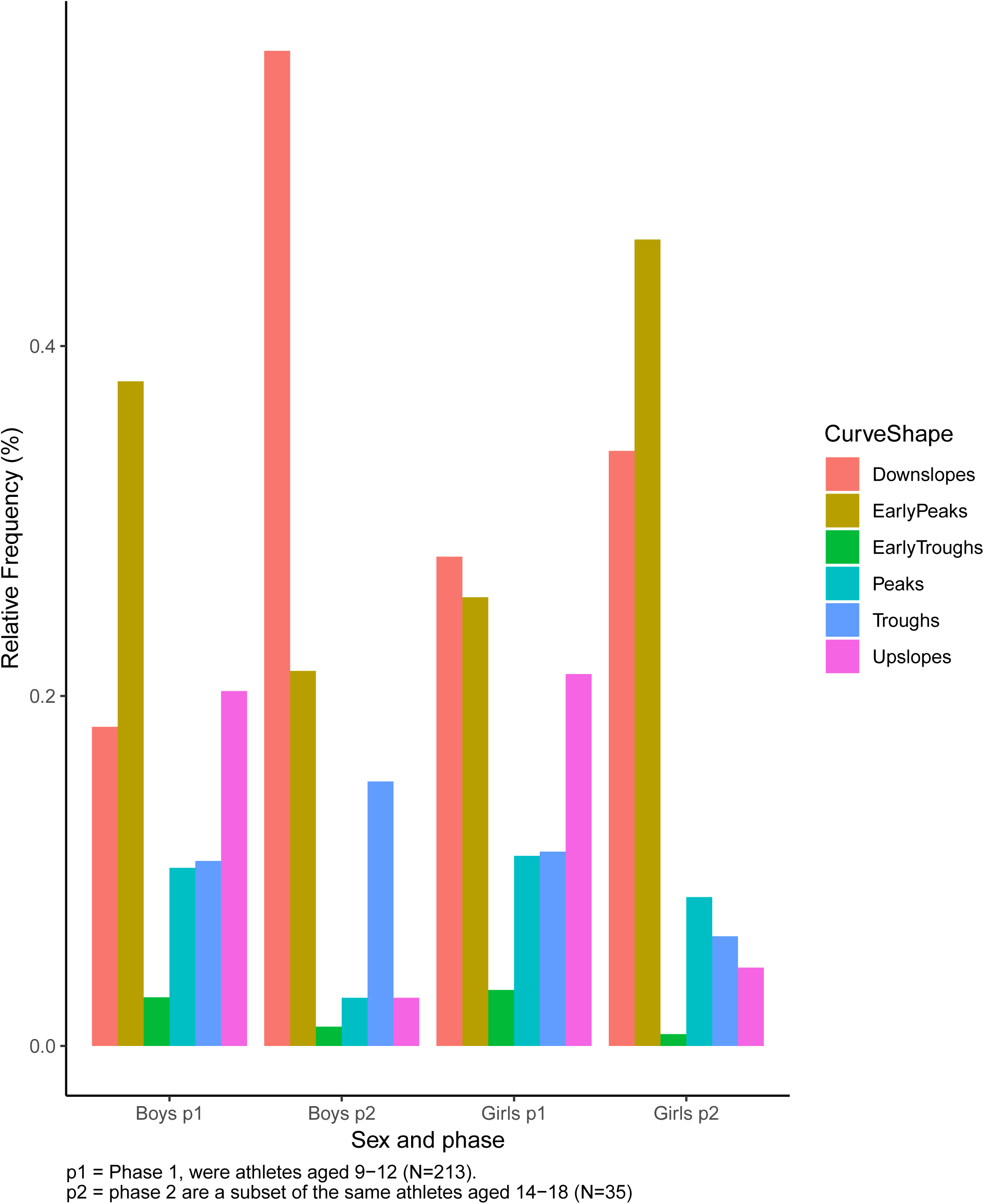
Relative frequencies of each shape according to sex and phase.

During further analysis of shape and magnitude based clusters, expected frequencies for early troughs and some groups of troughs were below 5 indicating that the assumptions of the Chi-square test are violated. A Monte-Carlo simulation procedure was used as a significance test (Adery 1968) instead. Analyses focusing on the knee valgus moment demonstrated that phase 2 boys had fewer than expected large early peaks, while phase 2 girls had the expected frequency (chi-square contributions of 17.1 and 1.4, respectively). For small early peaks, phase 2 boys had the expected frequency while phase 2 girls had higher than expected (chi-square contributions of 0.1 and 51, respectively). The total Chi-Square value of the test was 745 with P < 0.001. The observed and expected frequencies with chi-square contributions for shapes and magnitudes are reported in Table 3.

**Table 3:** Observed and expected frequencies for shape based clusters and magnitude based sub-clusters

|  |  | Observed (Expected) Frequencies |  |  |  | Chi-Square contributions |  |  |  |
| --- | --- | --- | --- | --- | --- | --- | --- | --- | --- |
|  |  | Boys |  | Girls |  | Boys |  | Girls |  |
| Curve shape | Magnitude classification | Phase 1 | Phase 2 | Phase 1 | Phase 2 | Phase 1 | Phase 2 | Phase 1 | Phase 2 |
| Early Peak | Small | 291<br>(261) | 58 (60) | 335<br>(422) | 130 (70) | 3.4 | 0.1 | 18.0 | 51.0 |
|  | Large | 284<br>(212) | 20 (49) | 307<br>(342) | 48 (57) | 24.8 | 17.1 | 3.5 | 1.4 |
| Peak | Small | 101<br>(112) | 10 (26) | 206<br>(182) | 33 (30) | 1.1 | 9.8 | 3.3 | 0.3 |
|  | Large | 53 (40) | 0 (9) | 66 (64) | 5 (11) | 4.4 | 9.2 | 0.0 | 3.0 |
| Downslopes | Small | 198<br>(268) | 83 (62) | 478<br>(434) | 77 (72) | 18.4 | 7.1 | 4.6 | 0.3 |
|  | Large | 78<br>(160) | 124 (37) | 220<br>(258) | 75 (43) | 41.7 | 205.8 | 5.5 | 24.1 |
| Upslopes | Small | 166<br>(167) | 7 (39) | 328<br>(269) | 18 (45) | 0.0 | 25.8 | 12.9 | 16.0 |
|  | Large | 139<br>(112) | 3 (26) | 204<br>(180) | 2 (30) | 6.7 | 20.2 | 3.1 | 26.1 |
| Troughs | Small | 114<br>(110) | 14 (25) | 199<br>(177) | 15 (29) | 0.2 | 5.1 | 2.6 | 7.1 |
|  | Medium | 43 (47) | 24 (11) | 70 (76) | 9 (13) | 0.3 | 16.0 | 0.4 | 1.0 |
|  | Large | 3 (11) | 17 (2) | 9 (17) | 4 (3) | 5.4 | 86.5 | 3.8 | 0.5 |
| Early Troughs | Small | 31 (33) | 3 (8) | 67 (54) | 3 (9) | 0.2 | 2.9 | 3.2 | 4.0 |
|  | Medium | 9 (7) | 1 (2) | 12 (11) | 0 (2) | 0.5 | 0.2 | 0.0 | 1.9 |
|  | Large | 2 (1) | 0 (0) | 1 (2) | 0 (0) | 1.1 | 0.2 | 0.2 | 0.3 |

The Chi-Square value is 745 and the P value of a Monte-Carlo simulation significance test is $< 0.001$

## Discussion

The main results of this study are in line with hypothesis 1a, i.e. that the two-step clustering process reported can differentiate between six different curve shapes of the knee valgus moment during the early stance phase, and 2-3 different magnitudes within each shape. Moreover, one of the shapes identified was the early peak, consistent with hypothesis 1b. In phase 1 boys had a higher relative frequency of early peaks, in contrast to hypothesis 2a. However, girls in phase 2 did have a higher relative frequency of early peaks with a ratio of 2:1, consistent with the reported 2-3x higher incidence of ACL injuries for adult females(Montalvo et al. 2018; Walden et al. 2011).

The incidence of female ACL injuries peaks in the late teens (Nicholls et al. 2017). Our phase 2 group has a mean age of 16 years. The relative incidence of ACL injuries in the amateur and pre-professional levels is higher than in the professional level (Montalvo et al. 2018), so our results of a 2:1 ratio is in the lower range. Keeping in mind that the injury mechanism involves multi-planar forces that include the knee valgus moment (Bates et al. 2018), this ratio is plausible. In contrast, research on the peak knee valgus moment for all of the weight acceptance phase has had inconsistent results of a fairly small magnitude (Sigward et al. 2012; Tanikawa et al. 2013) that is unlikely to explain a large difference in injury incidence.

Studies examining the effects of injury prevention programs with documented efficacy have failed to identify a plausible explanation (Thompson et al. 2017; Zebis et al. 2016) for why interventions can reduce ACL injuries by over 50% (Webster and Hewett 2018). Our results show that one potential target for intervention programs is to shift the curve shape of the early forces towards a pattern with no early peak. If the forces are delayed, it may provide sufficient time for protective muscles such as the hamstrings or hip abductors (Maniar et al. 2018) to reduce ACL load, or for the athlete to adopt a posture with greater knee flexion (Markolf et al. 1990). Future studies should examine the effects of injury prevention programs on the relative frequency of the early peak knee valgus moment shape.

The frequency of early peak knee valgus moments reported in this paper is not low enough compared to how rare ACL injuries are. However, the early knee valgus moment needs to be coupled with other factors in order to cause the multi-planar force event of an ACL rupture (Bates et al. 2018), notably the knee internal rotation moment. There is a uni-directional relationship between the knee internal rotation moment and the knee valgus rotations in cadavers (Kiapour et al. 2015). Similar variability in the shape of internal rotation moment curves may exist, and a co-occurrence of early peak knee valgus moment and knee internal rotation moments may be a rare event that resulting in high ACL loads and potential ACL fatigue (Lipps et al. 2013) or failure. Alternatively, a combination of high forces and an early peak may result in an injury. Our study is under-powered to detect whether the frequencies of small and large early peaks are correlated in phase 2 athletes.

The differences in shape between the six clusters are quite obvious when examined in the first 30% of the stance phase (Figures 2 & 3). However, when the whole stance phase curve is plotted for each shape (Figure 4), the visual differences between peaks and upslopes are minimal. What we call the peak shape has a peak occurring shortly before 30% of stance, while the upslope group has a local peak shortly after 30% of stance. Similarly, the whole-curve differences between downslopes and early peaks are small with only the initial 15% of stance being different. This could indicate that fewer than 6 movement strategies are used by the athletes, and differences in the initial 30% of stance may result from small variations in these movement patterns instead of from a separate movement strategy. Future studies should examine a potential relationship between athletes’ movement strategies during high-risk maneuvers such as a change of direction task, and the resulting valgus moment curve which may lead to identification of more specific targets for injury prevention programs.

### Strengths

This is to our knowledge the first study to have used cluster analysis techniques on 3D motion capture data. During motion capture, a number of different time series are calculated resulting in thousands of data points per measurement. Traditionally, these thousands have been reduced to a small number of single values such as local or global peaks (Leppanen et al. 2017; Torry et al. 2011) which can be input into a statistical model. Our results show that reducing different curve shapes to a single data point results in many of the data points being essentially an error due to a disconnect to the ACL injury mechanism, and thus reducing the statistical power of the study. We have further shown that techniques that make use of a greater amount of data can yield important insights and new avenues for future studies.

This study includes a large number of subjects with over 4900 repetitions of a change of direction task. A larger data set is necessary to ensure a cluster analysis process that is robust, but pre-testing with a small subset of data (not reported) has shown that the process is reproducible with a much smaller data set as well meaning the cluster analysis method can be readily used in smaller trials.

### Limitations

Assessment of homogeneity of shape within each cluster was performed via visual inspection. This requires a certain level of clinical judgement which may not be reliable between different assessors. The reader is encouraged to examine the results of the clustering process using our data and analysis script from the online depository [vantar link].

The C-Index of our cluster analysis is reported, but currently there is no consensus on what constitutes a good C-Index or how the number of clusters should be decided. We used the first elbow of the C-Index graph, or a 0.05 cut-off of a smooth graph. The potential number of clusters in our data is 1025 unique shapes, some of which likely differ only in the exact location of local maxima or minima. It’s possible, but in our opinion unlikely, that using 6 initial clusters would yield the same 6 shapes presented as the basic shapes in the present study, or that using 100 initial clusters would yield superior results.

The athletes were non-elite, while most video analyses of ACL injuries are performed in higher performance athletes(Hewett et al. 2009; Koga et al. 2018; Montgomery et al. 2018; Walden et al. 2015), likely because of higher availability of videotaped injuries. The decision to use the first 30% of the stance phase was made on the grounds that the fastest athletes in our cohort displayed an early peak at a timing consistent with ACL injury, and that time point was generally before 25%. A longer period (30%) was used to capture both sides of this peak to make a fully formed shape. The cut-off is likely to influence which trials fall in the upslope and which fall in the peak categories. However, as the early-peak occurs around or within 10% of the stance phase, we do not believe the choice of cut-off influence our results as reported.

## Conclusions and Clinical Relevance

This is to our knowledge the first study that demonstrates that clustering techniques are feasible to extract information from biomechanical data in a manner that can be done relative to a specific injury mechanism. A small number of distinct shapes of early stance phase knee valgus moment curves exist and can be identified with a cluster analysis procedure. The early peak knee valgus moment shape is consistent with the ACL injury mechanism, since the injury occurs early and the knee valgus moment can strain the ACL. The sex-specific frequencies of the early peak shape in adolescence is consistent with the sex-specific difference in ACL injury incidence and may be a predisposing factor to injury. These findings may lead to classification of movement strategies to identify neuromuscular factors associated with higher risk of ACL injury, thus improving the screening of athletes and content of injury prevention programs.

## Data Availability

A number of decisions have to be made by the authors during data processing and analysis. The field of cluster analysis has not reached consensus on how to select many important parameters during the cluster analysis process, including the number of clusters. In order to facilitate replication and auditing of the results presented, a data set that can be used to replicate the cluster analysis will be uploaded at datadryad.org should the manuscript be accepted. The data set includes the R code used to perform the cluster analysis. The data is uploaded as additional documents for submission in JEO.

## List of abbreviations

ACL: Anterior Cruciate Ligament

## Declerations

### Ethics

Ethics approval granted by the Icelandic National Bioethics committee, approval code VSNb2012020011/03.07. All participants, together with a legal guardian, received information regarding the study protocol, including known risks and benefits, and signed an informed consent statement.

### Funding

Funding for the study was provided by The Icelandic Centre for Research (Rannís – Rannis.is), funding codes 120410021, 903271305, 1203250031, and 185359051. The Football Association of Iceland has provided travel funds to the lead author to present findings at a conference. No funding source was involved in the study design, the execution of the study, the data analysis, writing the report, the decision to publish the results, or writing the article.

### Authors Contributions

HBS collected phase 2 data, designed and performed the data analysis and wrote the manuscript. KB designed the study, participated in planning of the data analysis, and provided substantial revision of the manuscript.

## Acknowledgements

The authors would like to acknowledge Prof. ólafur Pétur Pálsson and Ragnar Leví Guðmundarson for valuable input regarding the data analysis.

## Consent for publication

Not applicable.

